# Predicting Hypertension Among HIV Patients on Antiretroviral Therapy in Rural Eastern Cape, South Africa Using Machine Learning

**DOI:** 10.1101/2025.01.11.25320377

**Authors:** Urgent Tsuro, Trymore Ncube, Kelechi E. Oladimeji, Teke R. Apalata

## Abstract

**Background:** Hypertension continues to be a major challenge in developing countries like South Africa, as it significantly contributes to the cardiovascular disease burden in these countries. This study aimed to utilize the machine learning (ML) models to anticipate the incidence of hypertension in HIV patients under antiretroviral therapy (ART) in rural Eastern Cape, South Africa.

**Methods:** This research carried out a retrospective cohort study and created and tested six machine learning algorithms: Neural Networks, Random Forest, Logistic Regression, Naive Bayes, K-Nearest Neighbours and XGBoost. The goal was to predict the likelihood of developing hypertension. Feature selection was done using the Boruta method and the model was assessed using several metrics including aiming, precision, recall, F1 score, and area under the receiver operating characteristic curve (AUC).

**Results:** XGBoost outperformed all other models with an AUC of 0.96, which further suggests it can effectively distinguish between hypertensives and normotensives. In the case of Boruta analysis, some aggravated risk factors were age category, time on ART, BMI category, waist to hip ratio, waist size, family history of HBP and relationship status, physical activity, LDL cholesterol level, awareness of high blood pressure, education level, use of ART and diabetes mellitus.

**Conclusions:** This study has highlighted the utility of XGBoost, as one of the advanced machine learning algorithms, in reliably forecasting the occurrence of hypertension in HIV ART patients in a rural setting. The established risk factors elucidate the complexity behind the hypertension emergence and hence the need for triad approaches which include lifestyle changes, clinical treatments, and demographic solutions to tackle the public health problem.

## Introduction

Hypertension (HTN) is a global challenge, which is claiming approximately eight and half million individuals annually. Most of these deaths (88%) occur in low and middle-income countries (LMICs) [1]. Hypertension is also implicated in causing most cardiovascular disease (CVD) cases, particularly in South Africa where HTN prevalence is increasing at an alarming rate [1]. The increase in the prevalence of HTN is a result of different risk factors such as sedentary lifestyle and diet due to urbanization in South African societies [2]. Over the past thirty years South Africa also has shown a limited ability to detect, manage as well as treatment of HTN [3].

South Africa has low level of public awareness, health promotion and inadequate screening practices, which results in a significantly high percentage of undiagnosed HTN cases. Considering South Africa’s low level of public health awareness regarding HTN, the country’s HTN burden increases as untreated HTN can lead to severe health complications, such as stroke, kidney failure, heart failure, coronary artery disease, and premature mortality [4]. Nevertheless, it is crucial to note that these adverse outcomes are largely preventable through cost-effective and accessible treatments and interventions [5].

The high HTN incidence in South Africa is due to several factors such as limited health literacy, physical inactivity, unhealthy dietary habits, smoking, inadequate access to healthcare services, and the financial situation [2]. South Africa endures a dearth in reliable HTN research as most research often suffer from unbalanced samples, small sample sizes, and inconsistent methodologies for measuring risk factors. Furthermore, factors such as obesity, prolonged Human Immunodeficiency Virus (HIV) infection, diabetes, and aging are independently associated with an increased risk of developing HTN [6].

Hypertension affects people from all walks of life for a variety of reasons. Males often develop HTN at an earlier age than females [7]. According to Li (8), HTN is more common in older people, as they discovered that age is a crucial factor in HTN, with prevalence rates significantly increasing beyond the age of 45 [8]. According to Madela and Harriman (9), education level is a significant predictor of HTN, with lower levels related with higher HTN prevalence and higher levels with better health outcomes. Furthermore, unemployed people have higher stress levels and less access to healthcare than employed people [10], and Ramezankhani (11) found that married people have less HTN development than single, divorced, or widowed people, who are more vulnerable due to social isolation and stress.

Dietary habits have a substantial impact on HTN risk, with high salt intake being a well-established risk factor [12], and smoking has been linked to blood vessel damage, which raises the likelihood of developing HTN and thereby boosting CVD risks [13]. Furthermore, excessive alcohol consumption raises the incidence of HTN, although moderate consumption may have preventive effects [14]. Individuals who engage in regular physical activity, on the other hand, are less likely to develop HTN than those who lead sedentary lifestyles [15]. Anthropometric parameters such as Body Mass Index (BMI), which is classified as overweight or obese, are powerful predictors of HTN [16], and increasing waist circumference and waist-to-hip ratio are substantially related with increased HTN risk [17].

Medical conditions such as diabetes mellitus (DM) significantly increase HTN risk due to their impact on vascular health and metabolic function [18], alongside elevated glucose levels [19]. A family history of HTN also increases the risk of an individual to develop HTN [20]. Elevated cholesterol levels Low-Density Lipoprotein (LDL) cholesterol, specifically total cholesterol, triglycerides, and low High-Density Lipoprotein (HDL) cholesterol are major risk factors associated with the development of HTN [21]. Lower Estimated Glomerular Filtration Rate (eGFR) is associated with higher HTN risk as it signifies reduced kidney function [22]. Finally, chronic inflammation and metabolic changes associated with both HIV infection and antiretroviral therapy (ART) among people living with HIV (PLHIV), especially those on ART for longer duration correlates with increased HTN risk [6].

Risk assessment models are reported as being effective in identifying and classifying patients based on their risk factors, this facilitates the commencement of preventive methods. Notable examples include the Framingham Risk Score for predicting coronary heart disease [23] and the American College of Cardiology (ACC) / American Heart Association (AHA) Pooled Cohort Equations Risk Calculator [24]. However, these models often suffer from insufficient ethnic diversity, population representativeness, and limited reliability [25]. Hence, there is a persuasive need for developing risk prediction models specifically tailored to the rural populations in South Africa.

In terms of developing risk stratification tools for diagnosing HTN, machine learning (ML) techniques have recently proven to be superior to classic statistical methods [26]. Machine learning enables systems to learn and handle big datasets with complex interactions [27]. Machine learning also excels in estimating causal effects in observational research, while not being traditionally based on causal inference like statistical approaches [28]. Machine learning frequently outperforms traditional statistical methods by eliminating bias, automatically managing missing variables with minimal data manipulation, correcting for confounding factors, and balancing datasets, resulting in better outcomes [29]. Furthermore, ML algorithms excel at analysing large amounts of data, where traditional statistical methods may fall short [30], and provide precise measurements [31]. Therefore, ML technics can be useful in developing automated tools for disease prediction, decision support, and assessing HTN risk within a population [32].

A recent review highlighted ML approaches for detecting HTN and noted lack of studies that combine sociodemographic and clinical data with signal processing for enhancing model performance [29, 33]. One study employed ML algorithms to group HTN cases based on personal characteristics but did not consider sociodemographic data [34]. In South Africa, another study applied ML to DM and HTN risk stratification algorithms for 2,278 patients’ data captured by the community health workers [35]. Furthermore, ML methods employed for detecting HTN from electronic health records were also undertaken in South Africa [36]. However, while there is progress in ML models for individual disease risk prediction, there is no research that has been carried out predicting HTN risk factors among PLHIV in the rural Eastern Cape (EC), South Africa employing ML models.

The goal of this study was to create and compare a variety of prediction models for hypertension using different machine learning and statistical approaches in order to determine the most accurate model. In this case, the study aims at selecting the best model for real world use, while also determining the important factors related to PLHIV in rural EC, South Africa to improve insight against hypertension risk factors, hence the gaps in the study that is currently available.

## Methods

### Ethical Approval

This study was in line with the ethical guidelines as declared by Helsinki (37) and approval was obtained from Walter Sisulu University ethics committee, protocol number (048/ 2019) and the EC department of Health (EC_201907_020). Before completing a written informed consent, potential participants were issued with an information sheet in English and their vernacular. The information sheet contained the process of research, the rights of the participants, as well as the contact person’s information.

### Study Setting and Population

This study was conducted in the rural EC province of South Africa, which is an area heavily burdened by HIV. Thus, it was important to the effects of HIV and its treatment in this population hence, highlighting the intersection of healthcare limitations and chronic disease burdens.

The study population consisted of PLHIV who were on Highly Active Antiretroviral Therapy (HAART) and receiving medical care in healthcare facilities throughout rural EC. The current study involved PLHIV that were at least 18 years of age, which is ideal in capturing the sexually active age group [38]. To ascertain that the effect of HAART on hypertension in PLHIV is completely catered for, the participants were supposed to have been on HAART for at least 12 months. Also, as part of eligibility criteria, patients were expected to fulfil all the clinical requirements, including an in-depth history check, blood pressure assessment, and any other clinical history as deemed essential to guarantee reliable results and thorough analysis.

We determined precise criteria for inclusion and exclusion to enhance the concentration of the research. Participants comprised adult PLHIV patients who had initiated HAART for at least one year and had thorough clinical information. We did not include patients who had a history of HTN during active HAART. This was to avoid other factors that would interfere with the clarity of the treatment of HIV relations history with HTN. Patients who lack vital clinical information or have inadequate medical history records were also excluded to complement the aim of the study to enhance only robust and comprehensive datasets for prediction modeling and analysis.

### Study Design

A retrospective cohort study using ML techniques to model the predictors of incidence of HTN using data collected from selected health facilities in rural EC, South Africa.

### Data Analysis

As part of data analysis, numeric data were noted as mean ± standard deviation (SD) and for all categorical variable frequency and percentage were recorded. To assess the differences between hypertensive and normotensive groups, a chi-square test was utilised and a p-value < 0.05 was regarded as statistically significant.

We then investigated the six most used supervised ML models to measure their predictive effectiveness in diagnosing HTN. In this study, the development of HTN was considered as a target feature while the other features were considered as independent variables. Feature selection was performed using the Boruta based feature selection [39]. Following model training, we measured the accuracy, precision, recall, F1 and area under the curve (AUC). The higher accuracy and precision suggested comparatively better models. In addition, we conducted feature ranking, which highlighted factors that mostly contributed to the development of HTN. Model building and data analyses were performed using R studio, utilising packages like caret, ggplot, and tidyverse.

### Machine learning Models

Six ML models were developed which are, Logistic Regression (LR), Neural Network (NN), Naïve Bayes (NB), Random Forest (RF), k-Nearest Neighbour (kNN), and Extreme Gradient Boosting (XGBoost), these ML models were used to predict the development of HTN among PLHIV in rural Eastern Cape.

### Logistic Regression

Logistic regression is a fundamental ML model widely used for binary classification tasks. It can also be extended to handle multi-label classification problems [40]. The technique employs the sigmoid function to create a regression model that estimates the probability that an input belongs to a specific category [41]. Logistic regression is a binary classifier that takes one or more features as input and predicts the corresponding response [42]. It is known for its simplicity and effectiveness in delivering probabilistic predictions [43]. The logit function can be presented as

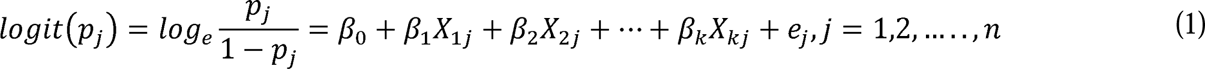

Where, *p_j_* is the probability of developing HTN and 1-*p_j_* is the probability of not developing HTN for *j^th^* individual and *β_k_* is the *k^th^* regression coefficient [44].

### Naive Bayes

Naive Bayes is a type of algorithm that belongs to the class of probabilistic types which uses the Baye’s theorem and assumes that there is no relation between the features in the layer [45]. This is a very basic but still effective classification approach based on Bayes theorem but with very strong independence assumptions [46]. The Naïve Bayes algorithm has a widespread application in many categories that include sentiment analysis, text classification, and medical diagnosis algorithms that presume that features are independent within a particular set [47]. Examples of where Naive Bayes has been used include assigning ratings in the airline business to customer satisfaction [48], text data in sentiment analysis [49] and military uses in denial-of-service attacks prediction [50].

### k-Nearest Neighbour

The kNN algorithm is a widely used classification approach in various fields. It relies on the idea that like objects are located near each other in a feature space, which is evaluated through distance triangulation [51]. In other words, the method defines the class as the mode of the classes of its k-nearest vertices, hence it is non-parametric is a learning method as well [52]. The kNN showed a promising performance in real-world settings and is thus often employed in supervised learning tasks [53].

### Extreme Gradient Boosting

XGBoost is an ML model which lies under decision tree-based ensemble methods that applies the Gradient Boosting framework, as explicated by Chen and Guestrin (54). This algorithm increases the accuracy of prediction models through sequentially applying new models of Decision Tree which corrects the failures of the previous models otherwise termed as sequential boosting [55,56]. XGBoost is highly efficient since it was developed for speed and performance, hence its popularity amongst the ML practitioners [57].

### Random Forest

Random Forest is an ensemble collection of models based on multiple Decision Trees, whereby each model can independently make predictions, and the outputs are aggregated through a voting process, using most of the models’ predictions [58]. Using this technique, the RF model can improve its significantly classification accuracy with quite a high value [59]. The model induces randomness into the tree construction which improves performance reliability. [60].

### Artificial Neural Network

Artificial Neural Networks are models that are inspired by the human brain, and they are also computational [44]. Among the various types of NNs, Multilayer Perceptrons (MLPs) are feedforward neural networks that consist of numerous layers which are input, output and hidden layers [61]. The input layer receives signals, the output layer provides classifications or predictions, and there can be multiple hidden layers in between, serving as the computational engine of the MLP [62]. Non-linear activation functions are typically used in both the hidden and output layers to introduce complexity and enable the network to learn complex patterns [63].

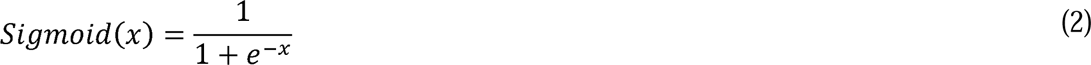

Where *x* is typically the linear combination of the input features and their corresponding weights, plus a bias term. Mathematically, it can be expressed as:

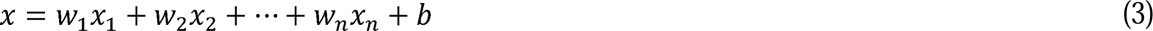

Where *x_i_* are the input features, *w_i_* are the weights and *b* is the bias.

### Performance Metrics

The predictions of the models developed in this study can generate four possible outcomes: True Positive (TP), True Negative (TN), False Positive (FP) and False Negative (FN). Positive individuals in our study had HTN, whereas negative patients had normal blood pressure. TP and TN are correct predictions. FP outcomes are positive predictions when they are negative. On the other hand, FN outcomes are predictions that are negative when, they are positive. We evaluate the prediction models with the following performance metrics:

**Sensitivity or recall or TP rate**: This metric indicates the proportion of true positives predicted out of all positives in a dataset [64].

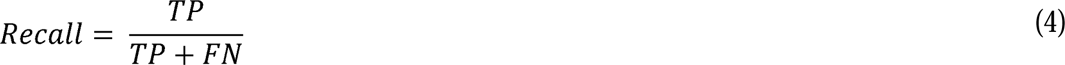

**Specificity or False Negative rate**: This is the number of negative cases that are mistakenly identified as positive [65].

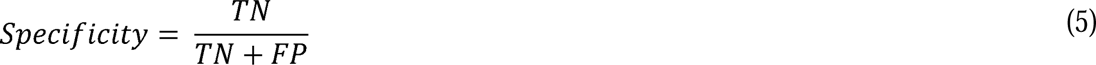

**Accuracy**: This is a metric utilised in determining how many correct predictions a model produced through the whole test dataset [66].

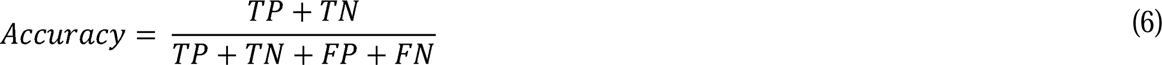

**Precision**: This indicates the correctness of the correct prediction [67].

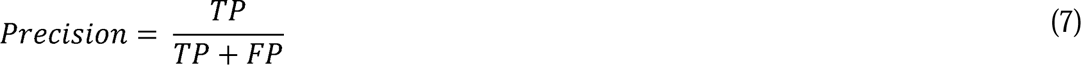

**F1-score**: This metric measures the accuracy of the model based on sensitivity and precision. A higher F1-score value indicates the model is more accurate [68].

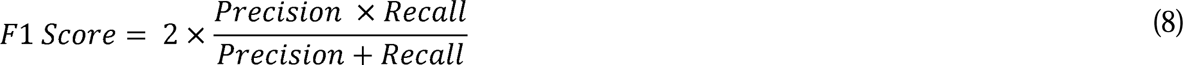

**Misclassification rate**: a performance indicator that indicates the proportion of incorrect predictions without differentiating between positive and negative predictions [69].

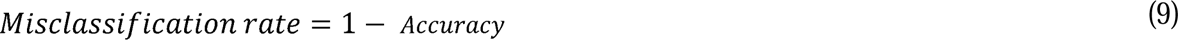

**Area Under the Receiver Operating Characteristic Curve** is used to assess the model’s prediction accuracy [44]. This statistic assesses the algorithm’s ability to distinguish between hypertensive and normal individuals [70].

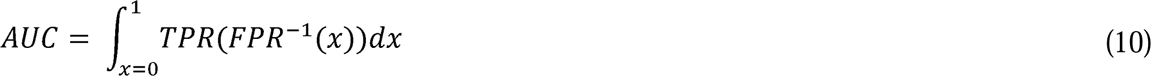

In scientific studies the true positive rate (TPR) which can be known as the sensitivity (y-axis) are plotted against 1-*specificity* (x-axis), which the false positive rate (FPR), this plot is known as the ROC curve. The ROC curve is essential in evaluating the predictive validity of ML-based model especially in the medical fraternity [44]. The AUC generated by a ROC has values from zero to one.

### Machine Learning Model Development Process

Fig 1 consists, a structured flowchart illustrating the comprehensive steps involved in the development of a ML model. This diagram serves as a visual guide through the sequential phases of a typical ML project, highlighting the critical stages and decisions made from data acquisition to the final evaluation and interpretation of the model.

**Fig 1.**
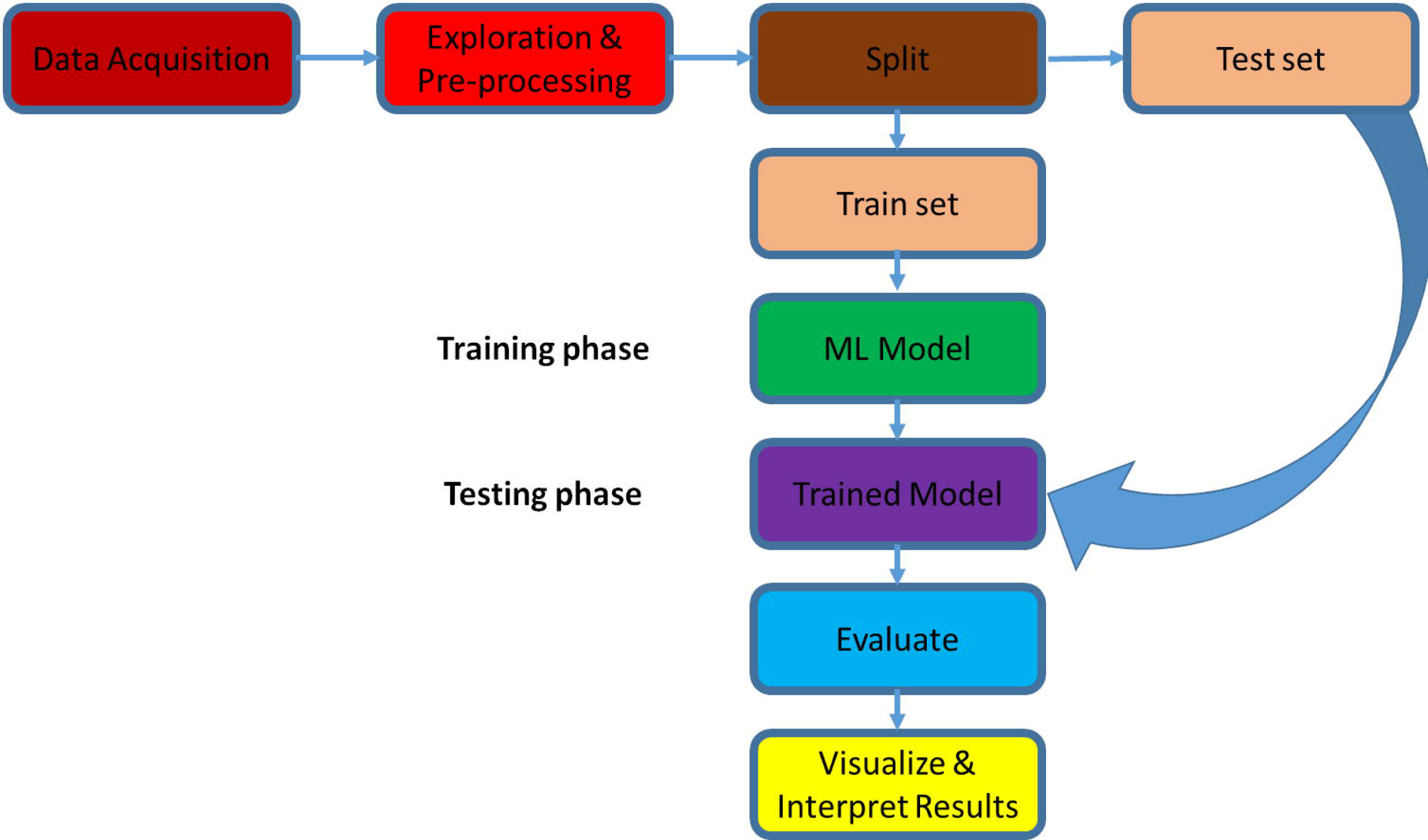
Machine Learning model for predicting HTN workflow.

The process commences with Data Acquisition, where the necessary data is gathered to form the foundation of the entire ML process. Exploratory analysis is performed on the data after data acquisition, followed by pre-processing of the data to identify patterns and anomalies. Data is then split into, training and testing sets in the ratio 80% and 20% respectively. Splitting the data is important so that we improve the quality of model output.

In the process of developing robust predictive models, it is crucial to ensure that the models perform well not only on the training data but also on unseen data to avoid overfitting [71]. To achieve this, the dataset was strategically split into training and testing sets in the ratio 80% to 20%, respectively. We compared the performance of the ML classifiers using accuracy, precision, recall and F-1 score respectively.

During the Training Phase, the training set is used to train the ML model, adjusting its parameters to best fit the data. The outcome of this phase is a trained model ready for testing and evaluation. In the Testing Phase, the trained model is evaluated using the test set to assess its performance and generalizability. The final step involves Visualizing & Interpreting Results, where the results are visualized, and the model’s performance is interpreted to make informed decisions or improvements.

This flowchart encapsulates the methodical approach in ML model development, providing a clear roadmap from data handling to obtaining a deployable model. The visualization aids in understanding the iterative nature of model training and the importance of each step in achieving a robust ML model.

## Results

### Sociodemographic characteristics of study participants

The results in Table 1 compromise an astute comparison between hypertensive and normotensive individuals, detailing on numerous lifestyle, demographic, and physiological characteristics. Most participants (73%) were female, and they had no significant difference in sex distribution between the normotensive (74%) and hypertensive (71%) groups with a p-value of 0.4. This finding suggests that sex is not a determinant of HTN prevalence within this cohort.

**Table 1.** Sociodemographic characteristics of study participants.

| Characteristic | N | Overall, N = 453 | Normotensive, N = 300 | Hypertensive, N = 153 | p-value |
| --- | --- | --- | --- | --- | --- |
| Sex | 453 |  |  |  | 0.4 |
| F |  | 331 (73%) | 203 (74%) | 128 (71%) |  |
| M |  | 122 (27%) | 70 (26%) | 52 (29%) |  |
| Age | 453 | 40 (30, 55) | 39 (31, 53) | 41 (28, 56) | 0.8 |
| Age Group | 453 |  |  |  | <0.001 |
| 18-25 |  | 45 (9.9%) | 38 (14%) | 7 (3.9%) |  |
| 25-35 |  | 70 (15%) | 64 (23%) | 6 (3.3%) |  |
| 35-45 |  | 113 (25%) | 62 (23%) | 51 (28%) |  |
| >45 |  | 225 (50%) | 109 (40%) | 116 (64%) |  |
| Education | 453 |  |  |  | <0.001 |
| None or Primary |  | 218 (48%) | 130 (48%) | 88 (49%) |  |
| Secondary or Higher |  | 235 (52%) | 143 (52%) | 92 (51%) |  |
| Employment | 453 |  |  |  | <0.001 |
| Employed |  | 217 (48%) | 138 (51%) | 79 (44%) |  |
| Unemployed |  | 236 (52%) | 135 (49%) | 101 (56%) |  |
| Marital Status | 453 |  |  |  | <0.001 |
| Single |  | 201 (44%) | 126 (46%) | 75 (42%) |  |
| Married |  | 96 (21%) | 60 (22%) | 36 (20%) |  |
| Widowed |  | 156 (34%) | 87 (32%) | 69 (38%) |  |
| Salt | 453 |  |  |  | <0.001 |
| Low |  | 129 (28%) | 85 (31%) | 44 (24%) |  |
| Moderate |  | 138 (30%) | 99 (36%) | 39 (22%) |  |
| High |  | 186 (41%) | 89 (33%) | 97 (54%) |  |
| Smoke | 453 | 235 (52%) | 144 (61%) | 91 (39%) | 0.6 |
| Alcohol | 453 | 229 (51%) | 133 (58%) | 96 (42%) | 0.3 |
| Exercise | 453 | 220 (49%) | 147 (67%) | 73 (33%) | 0.006 |
| BMI Category | 453 |  |  |  | <0.001 |
| Normal |  | 168 (37%) | 143 (52%) | 25 (14%) |  |
| Overweight |  | 1 (0.2%) | 1 (0.4%) | 0 (0%) |  |
| Obese1 |  | 25 (5.5%) | 9 (3.3%) | 16 (8.9%) |  |
| Obese2 |  | 90 (20%) | 46 (17%) | 44 (24%) |  |
| Obese3 |  | 169 (37%) | 74 (27%) | 95 (53%) |  |
| Tricep | 453 | 10 (9.00, 11.00) | 10 (9.00, 11.00) | 11 (9.00, 11.00) | 0.3 |
| Bicep | 453 | 11 (10.00, 12.00) | 11 (10.00, 12.00) | 11 (9.00, 12.00) | 0.3 |
| Subscapular | 453 | 13 (11.00, 15.00) | 13 (11.00, 15.00) | 13 (11.00, 14.00) | 0.9 |
| Suprailiac | 453 | 13 (11.00, 15.00) | 13 (11.00, 15.00) | 13 (12.00, 15.00) | 0.9 |
| WC | 453 | 112 (100, 127) | 113 (101, 127) | 111 (99, 126) | 0.5 |
| HC | 453 | 110 (105, 115) | 109 (105, 115) | 110 (105, 115) | 0.4 |
| WH Ratio | 453 | 1.02 (0.91, 1.15) | 0.94 (0.88, 1.02) | 1.23 (1.14, 1.29) | <0.001 |

Table 1 also shows that the median age of participants across all the groups was 40 years overall, of which 39 years in the normotensive group, and 41 years in the hypertensive group, with no significant difference p-value of 0.8. Nevertheless, when classified into age groups, there was a significant deviation (p < 0.001). The prevalence of HTN increased notably with age, with 64% of hypertensive PLHIV being over 45 years, compared to only 40% in the normotensive group. This indicates that older age is associated with a higher risk of HTN.

Employment status, though, showed statistical significance with a p-value that is less than 0.001, with 56% of hypertensive PLHIV reported as unemployed compared to 49% in the normotensive group, signifying a link between not being employed and chance of developing HTN.

Educational achievement seemed balanced generally, having 48% of participants with none or primary education and 52% in the secondary or higher education group. The highest level of education of the normotensive and hypertensive groups had a significant difference (p < 0.001), suggesting that education level might had a strong influence on HTN in this sample.

Both smoking group (p = 0.6) and alcohol (p = 0.3) consumption showed no statistical significance. A total of 235 PLHIV smoked and greater proportion (61%) were hypertensive while 49% were normotensive. and alcohol consumption at 49% and 53%, respectively (p = 0.3). However, physical exercise differed significantly (p = 0.006), with fewer hypertensive individuals (41%) engaging in regular physical activity compared to normotensive individuals (54%). This highlights the protective role of physical exercise against HTN.

Marital status was statistically significant differences with a p-value that was less than 0.001. The number of widowed PLHIV was greater for the hypertensive group (38%) compared to the normotensive group (32%). This might imply that social or emotional factors related to widowhood could influence HTN. The hypertensive group significantly too much salt (54%) than those who did not have hypertension (33%) and had a p-value < 0.001. This emphasizes the deep-rooted association between high salt intake and HTN.

Hip circumference (p = 0.4) and Waist circumference (p = 0.5) were found not being statistically significant for normotensive and hypertensive groups. Nonetheless, the waist-to-hip ratio was registered as significantly greater among the hypertensive PLHIV with a median of 1.23 compared to normotensive individuals, with a p-value <0.001. A greater proportion of those that were involved in some kind of physical exercise had less chance of developing hypertension (33%). This finding indicates that central obesity, as measured by the WH ratio, is a significant risk factor for HTN.

Body mass index (BMI) categories revealed significant differences (p < 0.001), with a greater proportion of obesity being among the hypertensive group. Specifically, 53% of hypertensive participants were Obese3 whereas 27% were in the normotensive group, highlighting a strong association between obesity and HTN. Skinfold measurements, including tricep, bicep, subscapular, and suprailiac values, showed little variation and no significant differences between groups, suggesting that these anthropometric measures are not strongly associated with HTN in this population.

In summary, the data highlight significant associations between HTN and factors such as employment status, age, salt intake, marital status, physical exercise, BMI, and WH ratio. These findings emphasize the complex, multifactorial nature of HTN and the importance of comprehensive lifestyle management in its prevention and control. Further research could explore causal relationships and the impact of targeted interventions to address these risk factors effectively.

### Health related characteristics of study participants

Table 2 provides a comparative analysis of several biochemical, lifestyle and health characteristics between the normotensives and hypertensives among PLHIV. The prevalence of DM is significantly higher in hypertensive individuals (66%), compared to normotensives (54%) with a p-value of 0.009. This highlights the common co-occurrence of HTN and diabetes, likely due to shared risk factors such as obesity and a sedentary lifestyle. In terms of glucose levels, there was no significant difference between the groups (p = 0.3), even though a slightly higher percentage of normotensive individuals had high glucose levels (42%) compared to hypertensive individuals (35%). This suggests that glucose levels alone do not significantly differentiate between hypertensive and normotensive states in this sample.

**Table 2.**
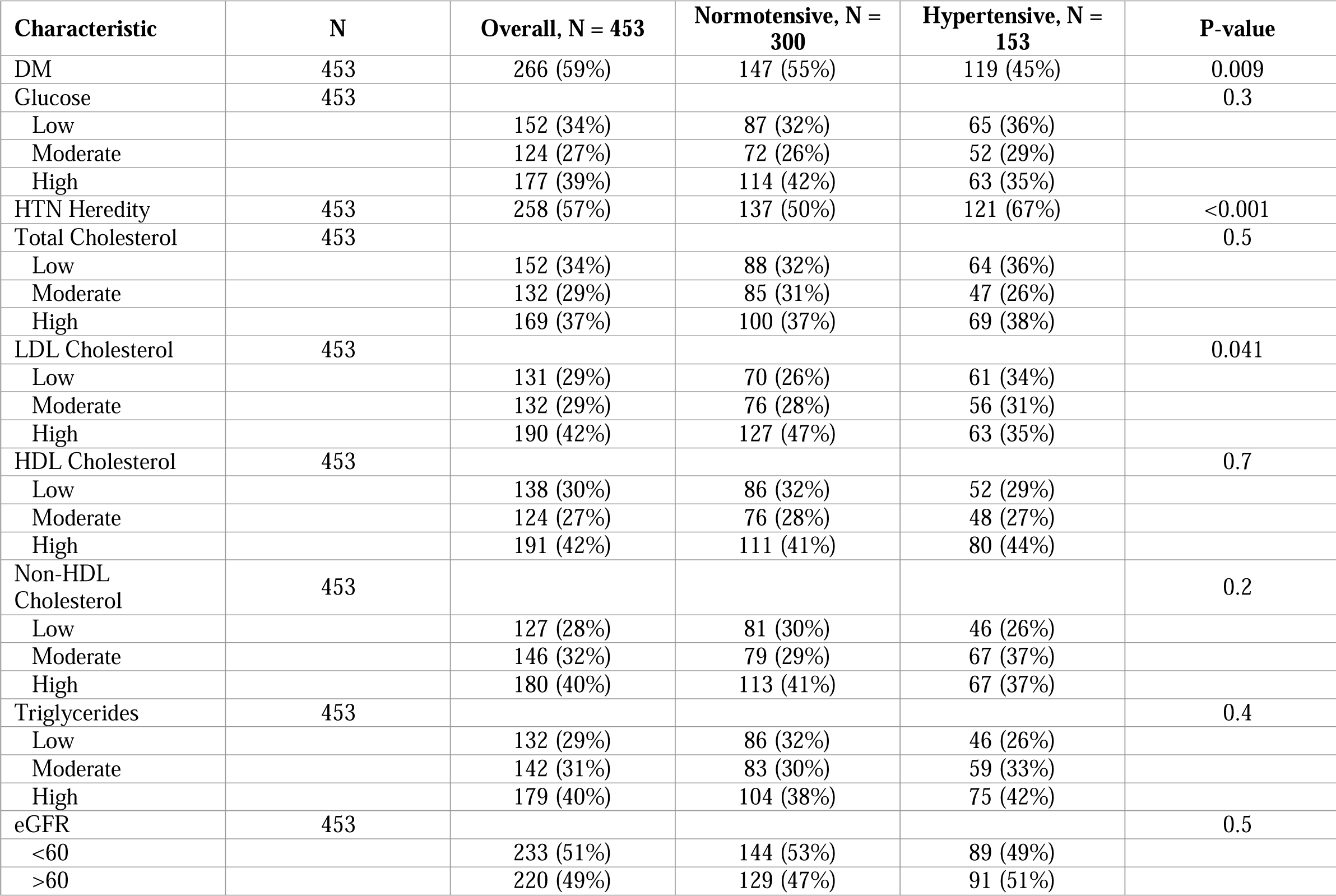
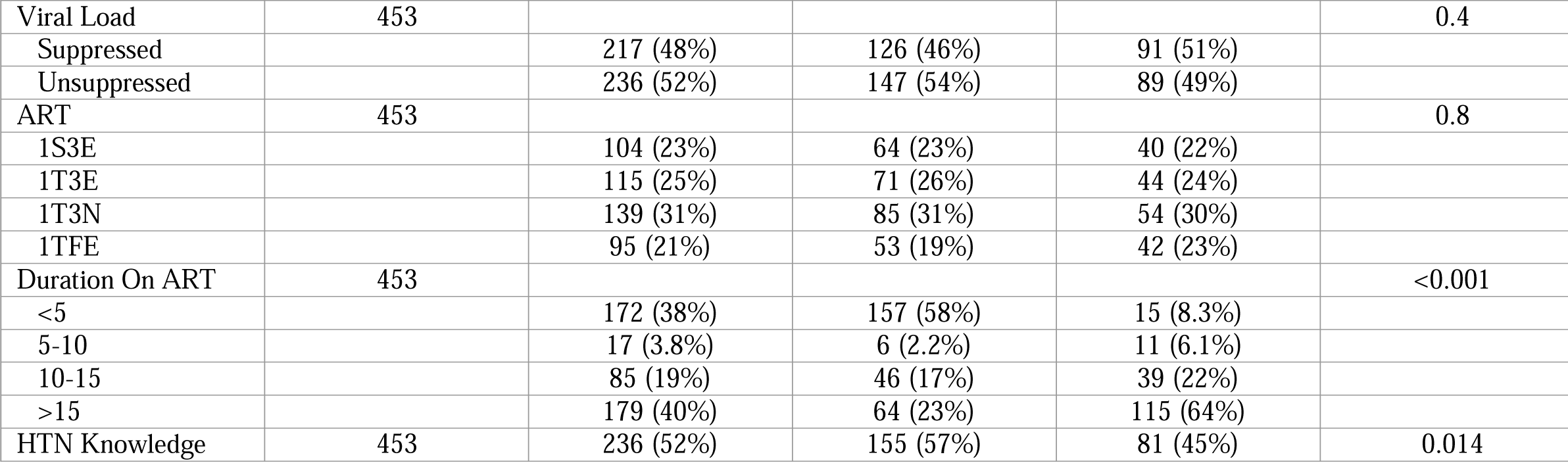
Health-related characteristics of study participants.

| Characteristic | N | Overall, N = 453 | Normotensive, N = 300 | Hypertensive, N = 153 | P-value |
| --- | --- | --- | --- | --- | --- |
| DM | 453 | 266 (59%) | 147 (55%) | 119 (45%) | 0.009 |
| Glucose | 453 |  |  |  | 0.3 |
| Low |  | 152 (34%) | 87 (32%) | 65 (36%) |  |
| Moderate |  | 124 (27%) | 72 (26%) | 52 (29%) |  |
| High |  | 177 (39%) | 114 (42%) | 63 (35%) |  |
| HTN Heredity | 453 | 258 (57%) | 137 (50%) | 121 (67%) | <0.001 |
| Total Cholesterol | 453 |  |  |  | 0.5 |
| Low |  | 152 (34%) | 88 (32%) | 64 (36%) |  |
| Moderate |  | 132 (29%) | 85 (31%) | 47 (26%) |  |
| High |  | 169 (37%) | 100 (37%) | 69 (38%) |  |
| LDL Cholesterol | 453 |  |  |  | 0.041 |
| Low |  | 131 (29%) | 70 (26%) | 61 (34%) |  |
| Moderate |  | 132 (29%) | 76 (28%) | 56 (31%) |  |
| High |  | 190 (42%) | 127 (47%) | 63 (35%) |  |
| HDL Cholesterol | 453 |  |  |  | 0.7 |
| Low |  | 138 (30%) | 86 (32%) | 52 (29%) |  |
| Moderate |  | 124 (27%) | 76 (28%) | 48 (27%) |  |
| High |  | 191 (42%) | 111 (41%) | 80 (44%) |  |
| Non-HDL Cholesterol | 453 |  |  |  | 0.2 |
| Low |  | 127 (28%) | 81 (30%) | 46 (26%) |  |
| Moderate |  | 146 (32%) | 79 (29%) | 67 (37%) |  |
| High |  | 180 (40%) | 113 (41%) | 67 (37%) |  |
| Triglycerides | 453 |  |  |  | 0.4 |
| Low |  | 132 (29%) | 86 (32%) | 46 (26%) |  |
| Moderate |  | 142 (31%) | 83 (30%) | 59 (33%) |  |
| High |  | 179 (40%) | 104 (38%) | 75 (42%) |  |
| eGFR | 453 |  |  |  | 0.5 |
| <60 |  | 233 (51%) | 144 (53%) | 89 (49%) |  |
| >60 |  | 220 (49%) | 129 (47%) | 91 (51%) |  |
| Viral Load | 453 |  |  |  | 0.4 |
| Suppressed |  | 217 (48%) | 126 (46%) | 91 (51%) |  |
| Unsuppressed |  | 236 (52%) | 147 (54%) | 89 (49%) |  |
| ART | 453 |  |  |  | 0.8 |
| 1S3E |  | 104 (23%) | 64 (23%) | 40 (22%) |  |
| 1T3E |  | 115 (25%) | 71 (26%) | 44 (24%) |  |
| 1T3N |  | 139 (31%) | 85 (31%) | 54 (30%) |  |
| 1TFE |  | 95 (21%) | 53 (19%) | 42 (23%) |  |
| Duration On ART | 453 |  |  |  | <0.001 |
| <5 |  | 172 (38%) | 157 (58%) | 15 (8.3%) |  |
| 5-10 |  | 17 (3.8%) | 6 (2.2%) | 11 (6.1%) |  |
| 10-15 |  | 85 (19%) | 46 (17%) | 39 (22%) |  |
| >15 |  | 179 (40%) | 64 (23%) | 115 (64%) |  |
| HTN Knowledge | 453 | 236 (52%) | 155 (57%) | 81 (45%) | 0.014 |

It should be noted that hypertension and diabetes often occur in the same individuals, which is most likely the result of excessive weight and physical inactivity. Comparing the groups in terms of glucose levels showed no major differences (p = 0.3), while the normotensive patients did display a higher proportion of high glucose levels at (42%) in comparison to the hypertensive patients who had it at (35%). It implies that glucose levels may not be the most effective indicators to distinguish between hypertensive and normotensive conditions in this group of patients.

In terms of HTN genetic factors there was a clear difference with 67% in hypertensive patients and 50% in normotensive patients having a family background for the condition (p < 0.001). It illustrates the potential physiological mechanisms through which genetic predisposition is linked to the HTN condition and provides insight into exposure prevention strategies among polygenic families. There were no differences between the groups with respect to the levels of total cholesterol (p = 0.5), which suggests that total cholesterol is not a significant differentiator of HTN in this sample. However, there was a significant difference in LDL cholesterol levels where significantly more individuals with normotension had high levels (47%) compared to individuals with hypertension (35%) with a p value of 0.041. This result reveals that there may be other variables responsible.

High-Density Lipoprotein cholesterol levels did not show statistically significant differences between normotensive and hypertensive individuals (p = 0.7) and the distribution of low, moderate and high HDL cholesterol was similar in both groups. This implies that HDL cholesterol might not be important in differentiating normotensive from hypertensive individuals. Non-HDL cholesterol levels were also found not to differ significantly between the groups (p = 0.2), suggesting that higher levels of non-HDL cholesterol by itself is not significantly associated with hypertension on this sample. The triglyceride levels were similar for both groups while differences between groups were not significant (p = 0.4) which also explains why triglycerides are not expected to play an important role in differentiating hypertensive status overall in this sample.

The estimated glomerular filtration rate (eGFR) values were also not significantly different in hypertensive individuals in comparison to normotensive individuals (p = 0.5) suggesting that kidney function, as determined by eGFR, is roughly the same in both groups, though renal failure is prevalent in patients with HTN. The viral load status of subjects, designated as suppressed and unsuppressed or active, did not differ significantly between the normotensive and hypertensive populations either (p = 0.4). The fact that the viral load status is similar in both groups indicates that it is not an important determinant of hypertension in this population.

The distribution of antiretroviral therapy (ART) regimens was consistent between normotensive and hypertensive individuals, with no significant differences (p = 0.8). This implies that the type of ART regimen does not significantly influence HTN status among participants. However, the duration on ART showed a significant difference (p < 0.001), with a higher proportion of hypertensive individuals being on ART for more than 15 years (64%) compared to normotensive individuals (23%). Conversely, a larger proportion of normotensive individuals had been on ART for less than 5 years (58%) compared to hypertensive individuals (8.3%), suggesting that longer duration on ART may be associated with an increased prevalence of HTN, possibly due to chronic exposure to ART-related metabolic effects.

Knowledge about HTN was also significantly different between the groups (p = 0.014), with a higher proportion of normotensive individuals demonstrating HTN knowledge (57%) compared to hypertensive individuals (45%). This suggests that increased awareness and knowledge about HTN may be associated with better blood pressure control and lower prevalence of HTN. Overall, the analysis highlights significant associations between HTN and factors such as diabetes mellitus, heredity, LDL cholesterol levels, duration on ART, and knowledge about HTN, underscoring the complex interplay of genetic, metabolic, and educational factors in the development and management of HTN.

### Data Partitioning

In ML, the exercise of dividing the dataset into ‘training’ and ‘testing’ sets is one of the primary actions which help to assess the performance and generalization of the models. The Table 3 describes the composition of two groups, hypertensive and normotensive, from the sample used for ML modeling. The dataset consists of two components; the first part is referred to as the Training Set which is used together with the data to train the ML models, and the test is the second part which is used for testing the performance of trained models with data that is not seen before to the neural networks. This approach avoids a situation where there is an overestimation of the performance of the models due to improper techniques in practice.

**Table 3.** Distribution of hypertensive and normotensive participants in the ML modelling dataset.

| Category | Training | Testing |
| --- | --- | --- |
| Hypertensive | 144 (39.7%) | 36 (40.0%) |
| Normotensive | 219 (60.3%) | 54 (60.0%) |

The participants’ distribution in the training and testing subsets is as follows: for hypertensive participants, the number was 144 (39.7 % of training set) and 36 (40.0 %) for test samples. For normotensive, the number stood at 219 (60.3%) for test samples and 54 for (60.0 % of the training set). It does so while ensuring that the distribution of the two sets of data across the set remains almost constant; in other words, the training and testing sets are representative of the entire data sample used.

It is key for the performance of machine learning models to be evaluated over representative samples so that they are built with a diverse range of categories for both the train and test sets in consideration. Such a practice greatly minimizes bias in the results of the models and ensures that they are both trained on and validated through an adequate sample of data.

### Feature Importance and Correlation Analysis of HTN Predictors

Figure 2A depicts a correlation and feature importance analysis graph affiliated with HTN Predictors. A closer view into the graph displays a rather distinct observation where the duration on ART seems to be the primary factor whereas other considerations greatly fall behind in relative importance ranking. This highlights the overdue influence that a sustained ART has on a patient where the healthy impacts are offset by long term metabolic impacts and the chances of developing chronic illnesses over time. Age group on the other hand comes in as a factor of equal importance as with age comes a plethora of chronic ailments. Increasing chances of suffering from hypertension or diabetes due to obesity also puts in body mass index as a significant modulatory factor.

A deep learning approach towards forecasting health outcomes treating body fat distribution metrics such as waist circumference and waist hip ratio as surrogate markers for central obesity has also proved to be quite efficient.

Salt intake, cardiovascular diseases, and genetics are on the important list in recognition of their association with high blood pressure or HTN. Other social determinants such as marital status, how regularly an opponent engages in physical activity, and how much low-density lipoproteins - LDL cholesterol - are in a person’s blood are health determinants as well. As does the knowledge and education one has about hypertension, as this chronic disease can be managed or avoided. Factors that can affect the person in question include employment status, diabetes mellitus- DM, and the use of antiretroviral treatment-ART, but these factors have a smaller effect when compared to primary determinants.

The interrelations of the vast array of variables under study are graphically depicted in a correlation matrix of the output in Fig 2B, the correlation matrix shows the correlation between two variables. It is logical that age group and duration of ART are positively related, meaning that older people are living longer and being treated for chronic diseases, as expected, for example, cancer. It also shows that waist to hip ratio correlates with BMI but does not rat control nor obesity. Correlation between diabetes and HTN predisposes each other which is one of the moderate relations.

**Fig 2.**
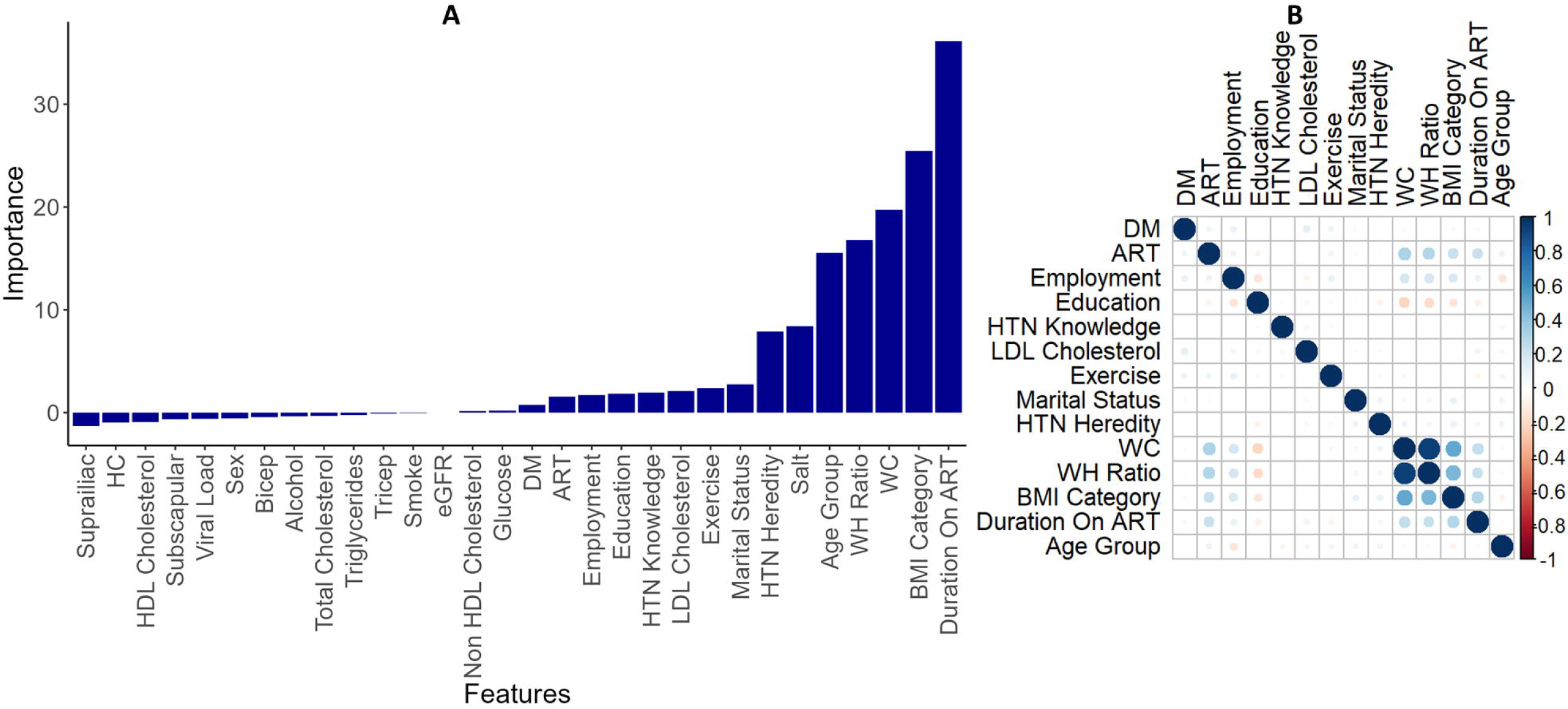
Risk factors selection. (A) Feature importance using Boruta, (B) Checking for multicollinearity among the selected features

Education and employment status are positively correlated, highlighting the socioeconomic link between educational attainment and job opportunities, which can significantly affect health outcomes. Regular exercise shows a positive correlation with lower LDL cholesterol levels, emphasizing the cardiovascular benefits of physical activity. Additionally, the relationship between marital status and HTN knowledge suggests that marital status may influence health literacy and access to health information, potentially due to better social support systems among married individuals.

The strong correlations between waist circumference, waist-hip ratio, and BMI category further highlight the relatedness of different obesity metrics, emphasizing the importance of addressing obesity in health interventions. Glucose levels are positively correlated with diabetes mellitus status, as expected, given that elevated glucose is a diagnostic criterion for diabetes. Lastly, the correlation between ART usage and viral load underlines the role of ART in managing viral infections, likely in the context of HIV/AIDS, highlighting the importance of effective treatment adherence in achieving better health outcomes.

In conclusion, these figures collectively emphasize the complex interplay between demographic, lifestyle, and clinical factors in determining health outcomes. The prominent roles of ART duration, age, BMI, and central obesity measures point to the need for comprehensive interventions targeting these areas to manage and prevent chronic diseases effectively. The correlations among these factors further illustrate the multifaceted nature of health determinants, underscoring the necessity of an integrated approach in healthcare management to address the intertwined influences of genetics, behaviour, and social determinants.

### Evaluating Machine Learning Models

The ROC curves in Fig 3 A and B provide a comprehensive evaluation of various ML models used for binary classification tasks, specifically measuring their true positive rates against false positive rates across different thresholds. The AUC values for each model indicate the effectiveness of the models in distinguishing between the two classes.

**Fig 3.**
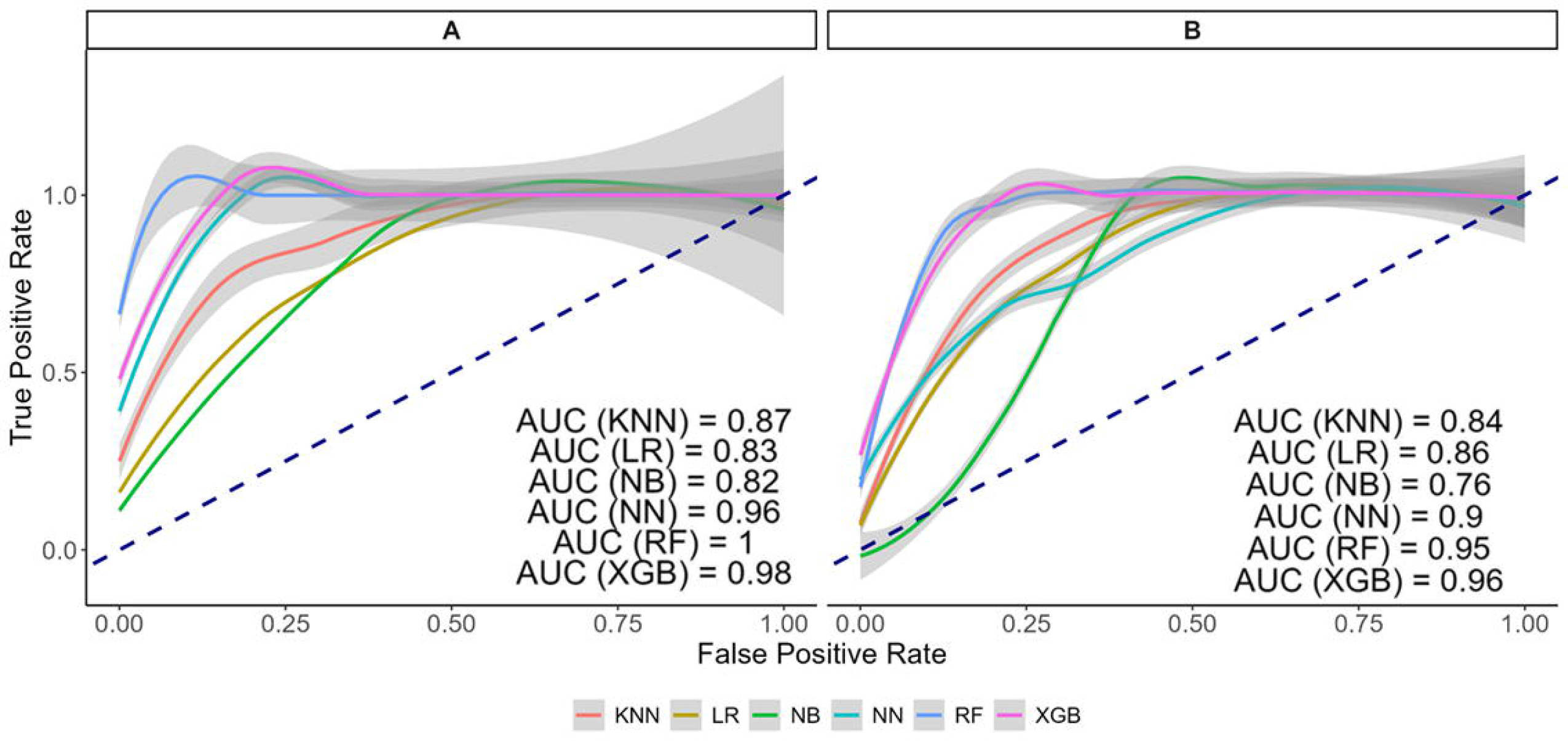
Receiver Operating Characteristic Curves. (A) Training set (B) Testing set

In Fig 3A, the ROC curves for the models KNN, LR, NB, NN, RF, and XGBoost are depicted. The Random Forest model stands out with an AUC of one, indicating perfect discrimination and suggesting that it can classify positive and negative cases without any errors. This could imply overfitting, where the model may perform exceptionally well on the training data but might not generalize as effectively to unseen data. The XGBoost model also performs exceptionally well, with an AUC of 0.98, highlighting its robust capability in classifying the data accurately. The Neural Network model follows closely with an AUC of 0.96, highlighting its effectiveness in learning and generalizing from complex data patterns.

The K-NN, LR, and NB models show lower AUC values of 0.87, 0.83, and 0.82, respectively. These models, while performing adequately, are less effective compared to the tree-based and neural network models. Their ROC curves approach the 45-degree diagonal line more closely, indicating a higher rate of false positives and less efficient classification performance, especially in more complex scenarios where capturing intricate data relationships is crucial.

In the current section, Fig 3B shows the models being evaluated once again, this time around the performance metrics lie in close similarity to each other but with slight variance. The Extreme Gradient Boosting model secures a reliable AUC of 0.96 for class distinction and thus proves to be strong. The performance metrics of the NN model are still significantly high with an AUC of 0.90, which is slightly lower than what was achieved in Fig 3A. This shows that the classification metrics are still consistent but with slight degradation. The same phenomenon is observed in RF model as well as it has further dropped to 0.95 from previously achieving a perfect classification score. While still being effective, this change suggests that performance may vary dependent upon the data subsets used or configuration of the model.

The performance of Scenarios in which the Naive Bayes’ model is placed in has improved as per the reading of section 3B which in turn has shifted the AUC value up to be 0.86. Such an increase is a sign of better classification than what was previously observed. Seeing a drop-off from AUC of 0.84 is the K-nearest neighbours model which does exhibit reasonable performance metrics, but they are outperforming by more advanced models. Even more at a disadvantage is the Naive Bayes model which has an AUC value of 0.76. The reason for this regression in performance is because the model can classify quite well due to the independent feature learning which could be obtained from complex datasets.

In conclusion, a comprehensive examination of Fig 3A and B provides a succinct summary of the merits and disadvantages of various ML models in single-valued classifications. Tree-based models such as Random Forest and Extreme Gradient Boosting continue to outrank others owing to their ability to account for non-linear relationships or interactions among the features. The Neural Network model also performs exceptionally well – showing little fluctuation over time due to its adaptability and learning from intricacies of multidimensional sets of data. Less complex models such as kNN, LR and NB, on the other hand, are relatively deficiently by way of complexity, and although they achieve moderate success, they are not as suited to intricacies in the data. This indicates that the model employed should be suitable for the data and the type of classification to be undertaken.

### Comparative Analysis of Machine Learning Models

In Table 4, the results analysis is provided covering accuracy, precision, recall, F1-score, AUC visualizing a comparison across multiple models kNN, Logistic Regression LR, NB, NN, RF and XGBoost. Out of the evaluated models, Random Forest and XGBoost turn out to be the best. Random Forest had the overall best performance which had an accuracy of 91.1% along with high precision (91.1%) and strong recall (94.4%) asserting a good case on its classification and positive case prediction ability. In a similar fashion, XGBoost had the same metrics for precision and recall (88.9%) and had a great AUC of 96.0% showing good accuracy on distinguishing ability. However, unlike the rest of the models, Naive Bayes had a high precision of 97.2% but a low recall of 64.8% showcasing being good at getting the positives right but forgetting some instances. Logistic Regression and Neural Network also had good performance as LR had a decent precision of 80.4% and recall of 83.3% while NN had a good AUC of 89.9% with good overall metrics. The need to personalize choice of models abiding by the accuracy, precision, recall, and general context specific predictive power of different practical contexts is the takeaway from the observations.

**Table 4.**
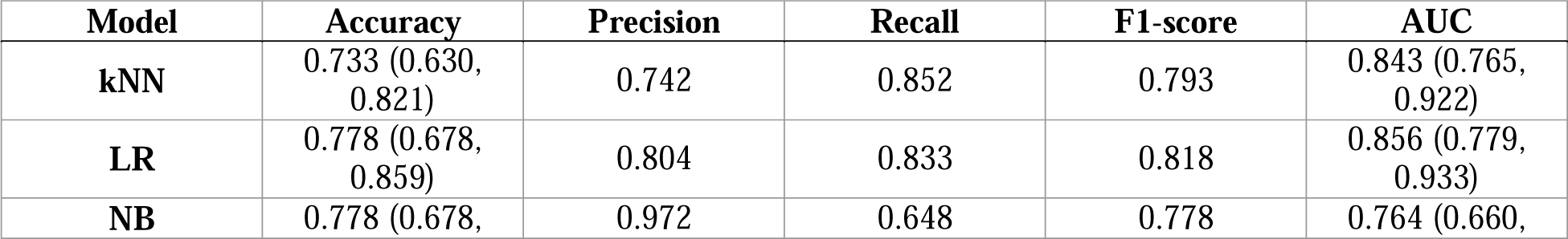
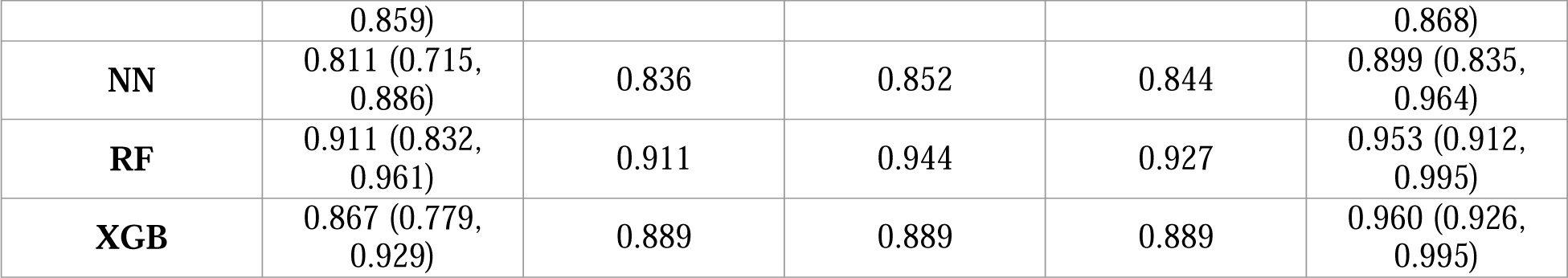
Comparative Analysis of ML Models.

### Performance and Generalization, Testing for Overfitting

Overfitting is one of the few phenomena that diminishes the performance of an ML-model. This is an aspect where a model learns the training dataset, and this then contributes towards the model’s tendency to predict on new data [72]. To further investigate if our models were over fitting, we compared the training testing accuracies our models produced.

From the performance metrics analysed, the bar chart for Fig 4 ML models comparison is detailed for several ML models. The models that were evaluated include LR, NN, kNN, NB, RF and XGBoost. The metrics assessed are Training Accuracy, Testing Accuracy, Accuracy Difference, Misclassification and AUC.

**Fig 4.**
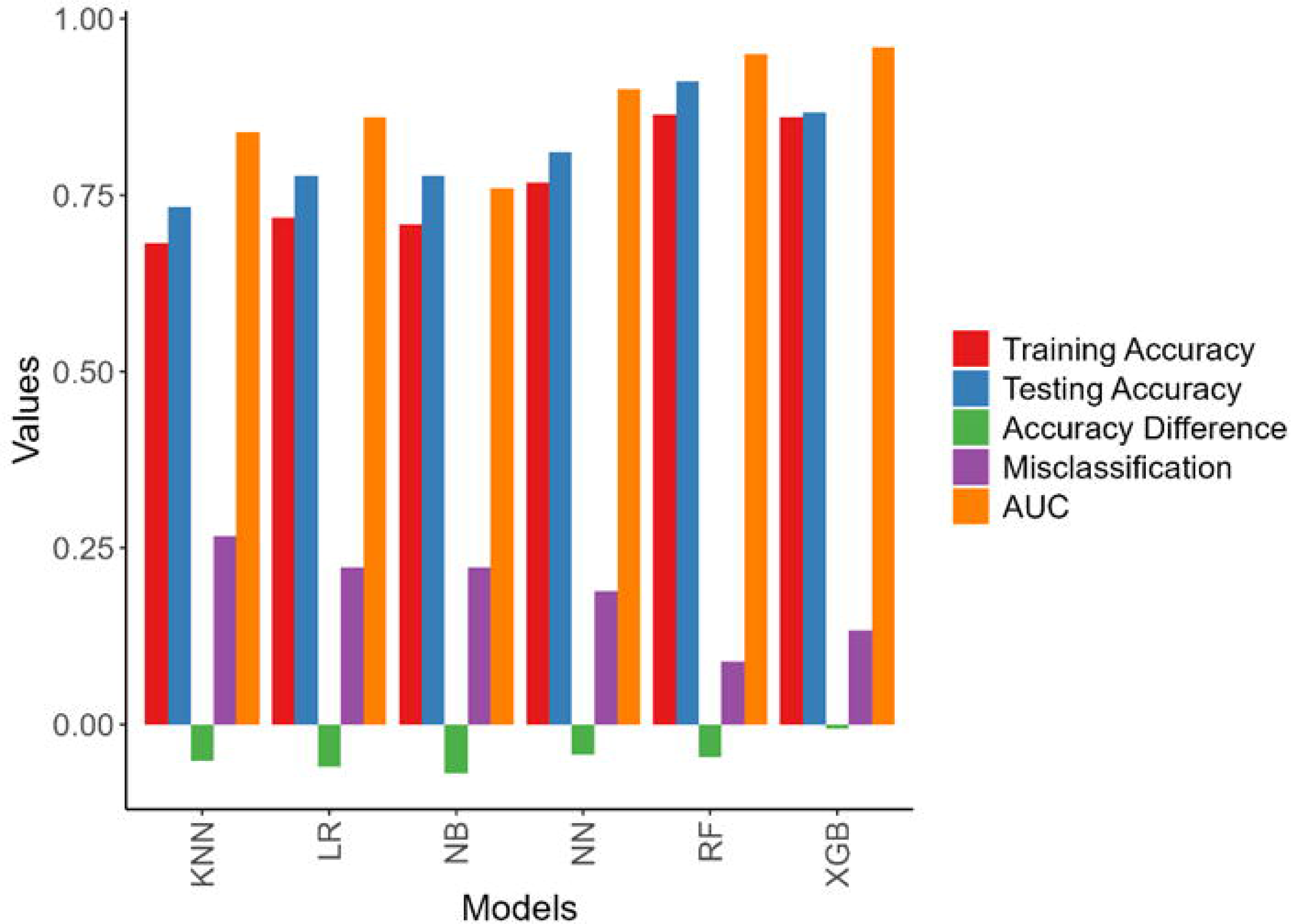
Bar plot to test under and over fitting.

It is clear from Fig 4 that Random Forest and XGBoost models achieves quite high training and testing accuracy, which shows the strength and efficiency of being able to deal with the dataset used. These models also have small accuracy difference for the cases of training and testing, which demonstrates that these models generalise well and are not over fitted. The AUC values for these models are also high which means that these models have good learning models.

On the other hand, the results obtained from the Naive Bayes model are not satisfactory, since it reports lower values in both training and testing accuracy while its misclassification rate is significantly higher. This may be due to its pronounced difficulties in dealing with the intricacies of the dataset or it is an under fitting problem. Overfitting problems are present in a Neural Network model which does not affect remarkably its accuracy metrics but generates a considerable accuracy gap still.

In terms of accuracy, KNN and Logistic Regression models perform moderately high however, they do not achieve the same effectiveness as the ensemble methods such as RF and XGBoost. Either the characteristics of the data or the capabilities of the model may account for this.

In conclusion, from the models developed in this analysis, RF and XGBoost are the best due to its comprehensive generalization and high performance. They score the highest among the models evaluated in terms of accuracy and generalisation performance. Nevertheless, XGBoost suffers from significant under fitting compared to RF so for this study XGBoost is the optimal choice.

## Discussion

In this study, we investigated numerous ML algorithms to propose an explainable framework for predicting the risk of HTN in rural EC, South Africa. We trained six ML algorithms (NN, RF, LR, NB, kNN and XGBoost) to predict HTN, utilising risk factors of HTN among PLHIV. The performance of the established models matched by precision, accuracy, F1-score, recall, and ROC curve with AUC value on testing set. Considering the performance measurements, we concluded that XGBoost is the most appropriate candidate classifier for predicting HTN in this population. This selection tallies with the selection made by Chowdhury, Leung (73) in a study conducted in Canada where they proposed a system on 18,322 individuals with 24 candidate risk factors.

A healthcare campaign began in 2022 which focused on identifying advanced machine learning techniques to determine hypertension. Throughout this campaign Oanh and Tung (74) were able to create an ML model which has the capability to forecast hypertension risk in patients stationed at Vietnam. To keep the models more advanced, algorithms such as KNN, SVM, NB, voting and boosting were utilized to increase versatility. Other metrics which were used during model testing included the likes of precision, recall and F1-score. A similar approach was done by Islam and Talukdar (32) during the final parts of their research project across India, Nepal and Bangladesh as well, which consisted of over 818000 participants. They looked at seven risk factors for HTN and tested several algorithms, including gradient trees, random forests, gradient boosting machines, XGBoost, logistic regression, and linear discriminant analysis. Just like the current study, they also found that XGBoost performed the best among the tested methods.

In Malaysia, Chai and his team (70) utilised data from 2,461 participants to build a system for diagnosing HTN. They applied three types of algorithms: neural networks, traditional models like logistic regression and decision trees, and ensemble models including random forests and Light gradient boosting machine (GBM). The models were evaluated for their ability to predict outcomes using metrics such as sensitivity, specificity, accuracy, and others, with LightGBM achieving the highest accuracy at 74.39%.

Islam et.al (76) used national data from Bangladesh, which included 6,965 participants and 13 risk factors for HTN. They then developed models using four ML algorithms and assessed their performance on a test dataset, using measures like accuracy, precision, recall, F1-score, and AUC. The gradient boosting model achieved the highest AUC score of 0.669.

Thus, the comparative results suggested that our proposed XGBoost framework could predict HTN with higher AUC. Moreover, Boruta analysis revealed that age group, duration on ART, BMI category, WH ratio, WC, HTN heredity, marital status, exercise, LDL cholesterol, knowledge of HTN, level of education, ART and DM were the important risk factors for developing HTN in the model.

A study by Belay and colleagues in 2022 (77), conducted in Ethiopia, revealed that individuals over 60 years old are twice as likely to develop HTN compared to those aged 18-40. This finding aligns with the conclusions of several other reviews and meta-analyses [78, 79]. The study highlights that aging is associated with changes in the arteries, particularly increasing stiffness in larger arteries, which contributes to the risk of HTN. Additionally, weight and body fat were identified as the second and third major factors driving HTN. Excess body weight, particularly visceral and retroperitoneal fat, plays a significant role in the development of HTN.

Another critical factor is BMI, which has been linked to HTN in earlier studies, including those by Hall et al. in 2019 (80). Body mass index may contribute to HTN and other cardiovascular issues by affecting the renin-angiotensin-aldosterone system and causing endothelial dysfunction, as noted by Imai in 2022 (81). Furthermore, high LDL was identified as a significant marker for HTN this supported the finding of a systematic review conducted by Obsa et.al (82). Additionally, other risk factors such as salt intake, alcohol consumption, and smoking were found to be important contributing risk factors of HTN, which is similar with other studies in literature [83, 84].

## Limitations

Although this work has many strengths, it also has some limitations; the study did not measure the quantity of alcohol, cigarettes, and salts that were consumed. More data is required to validate the findings of the study.

## Conclusion

This research delved into the incorporation of ML algorithms in predicting the incidence of hypertension among HIV infected patients receiving HAART in the healthcare of rural EC, South Africa. For building a useful and easy to understand framework for prediction, we trained and tested the models of six ML algorithms NN, RF, LR, NB, kNN, and XGBoost. According to our experiments, the XGBoost model produced the highest AUC score, and minimum overfitting compared to other models, thus the XGBoost model outperformed the other models. This suggested that XGBoost should be treated as the major classifier for HTN prediction in this situation. Our findings were consistent with previous studies supporting the fact that XGBoost as a reliable model for predictive analysis in an array of healthcare settings [73, 75, 70]. As highlighted by Boruta analysis, factors such as age group, duration on ART, BMI category, WH ratio, WC, presence of a HTN history in the family, marital status, physical activity, LDL cholesterol level, knowledge of HTN, level of education, ART treatment history, and diabetes incidence were identified as risk factors. All other factors improve the overall predictive accuracy of the model and illustrate the complexity of the risk factors for hypertension among PLHIV in rural settings of Eastern Cape, South Africa.

## Supporting information

Department of health clearance

Institutional clearance

## Data Availability

All data produced in the present study are available upon reasonable request to the authors

## Acknowledgments

We express our sincere gratitude to the Walter Sisulu University Research Unit, and we also thank the staff of the participating CHCs and clinics for their assistance. Most of all, we thank the patients who participated in the study for their full co-operation and trust.

## Author Contributions

**Conceptualization:** Urgent Tsuro, Trymore Ncube, Kelechi E. Oladimeji, Teke R. Apalata.

**Methodology:** Urgent Tsuro, Trymore Ncube.

**Investigation:** Urgent Tsuro.

**Data curation:** Urgent Tsuro.

**Software:** Urgent Tsuro.

**Visualization:** Urgent Tsuro.

**Formal analysis:** Urgent Tsuro.

**Validation:** Urgent Tsuro, Trymore Ncube.

**Writing original draft:** Urgent Tsuro.

**Writing review and editing** Urgent Tsuro, Trymore Ncube, Kelechi E. Oladimeji, Teke R. Apalata.

**Resources:** Teke R. Apalata.

**Supervision:** Teke R. Apalata, Kelechi E. Oladimeji.

## Funding

This research was fully funded by the South African Medical Research Council under its Research Capacity Development Grant (MRC-RFA-CC 01-2014). The funders had no role in study design, data collection and analysis, decision to publish, or preparation of the manuscript.

Institutional Review Board Statement: Walter Sisulu University’s Human Research Ethics and Biosafety Committee provided ethical approval (048/2019) and the Eastern Cape Department of Health (EC_201907_020) in accordance with the guidelines of the Declaration of Helsinki.

Informed Consent Statement: Consent forms were signed to obtain written informed consent for study participation. Personal identification information was removed from the data, and unique study codes were used to ensure participant anonymity.

Data Availability Statement Data will be made available upon request from the corresponding author.

