## Supplementary material for "Predicting Hypertension Among HIV Patients on Antiretroviral Therapy in Rural Eastern Cape, South Africa Using Machine Learning": Department of health clearance

Province of the
EASTERN CAPE
HEALTH

Enquiries: Yvonne Gixela Tel no: 079 074 0859

**Date: 07 June 2021**

**RE: Modelling the Incidence of Cardiovascular Diseases among HIV infected Patients receiving HAART in Rural Eastern Cape, South Africa. (EC_201907_020)**

**Dear Mr U. Tsuro**

The department would like to inform you that your application for the abovementioned research topic has been approved based on the following conditions:

1. During your study, you will follow the submitted protocol with ethical approval and can only deviate from it after having a written approval from the Department of Health in writing.

2. You are advised to ensure, observe and respect the rights and culture of your research participants and maintain confidentiality of their identities and shall remove or not collect any information which can be used to link the participants.

3. The Department of Health expects you to provide a progress update on your study every 3 months (from date you received this letter) in writing.

4. At the end of your study, you will be expected to send a full written report with your findings and implementable recommendations to the Eastern Cape Health Research Committee secretariat. You may also be invited to the department to come and present your research findings with your implementable recommendations.

5. Your results on the Eastern Cape will not be presented anywhere unless you have shared them with the Department of Health as indicated above.

Your compliance in this regard will be highly appreciated.

**SECRETARIAT: EASTERN CAPE HEALTH RESEARCH COMMITTEE**
