## Supplementary material for "Predicting Hypertension Among HIV Patients on Antiretroviral Therapy in Rural Eastern Cape, South Africa Using Machine Learning": Institutional clearance

WSU
Walter Sisulu University

FACULTY OF HEALTH SCIENCES

POSTGRADUATE EDUCATION, TRAINING, RESEARCH AND ETHICS UNIT

**HUMAN RESEARCH COMMITTEE**

**CLEARANCE CERTIFICATE**

| | |
|---|---|
| PROTOCOL NUMBER | : 048/ 2019 |
| PROJECT | : MODELLING THE INCIDENCE OF CARDIO-VASCULAR DISEASES AMONG HIV INFECTED PATIENTS RECEIVING HAART IN RURAL EASTERN CAPE. SOUTH AFRICA |
| INVESTIGATOR(S) | : MR TSURO URGENT |
| DEPARTMENT | : PUBLIC HEALTH |
| DATE CONSIDERED | : 26 JUNE 2019 |
| DECISION OF THE COMMITTEE | : APPROVED |

**N:B** You are required to provide the committee with a progress or outcome report of the research after every 6 months. The committee expects a report on any changes in the protocol as well as any untoward events that may occur at any time during the study as soon as they occur.

**WALTER SISULU UNIVERSITY**
**ACADEMIC HEALTH SERVICE COMPLEX OF THE EASTERN CAPE**
**POSTGRADUATE EDUCATION AND TRAINING**
**FACULTY OF HEALTH SCIENCES**
**WALTER SISULU UNIVERSITY**
**P/BAG X 1, WSU, 5117, E.C**

DR T APALATA
CHAIRPERSON

28-06-2019
DATE

**DECLARATION OF INVESTIGATOR(S)**

(To be completed in duplicate and one copy returned to the Research Officer at Office L311, 3rd Floor, Old Library Building, NMD Campus, WSU)

I/We fully understand the conditions under which I am/we are authorized to carry out the abovementioned research and I/we guarantee to ensure compliance with these conditions. Should any departure to be contemplated from the research procedure as approved I/we undertake to resubmit the protocol to the Research Ethics Committee. I/We agree to a completion of a yearly progress report.

24/07/2019

N. B. Please quote the protocol number in all enquiries.

Scanned by CamScanner
